# Evaluation of Vaccination Strategies for the metropolitan area of Madrid

**DOI:** 10.1101/2021.11.29.21267009

**Authors:** David E. Singh, Carmen Olmedo Lucerón, Aurora Limia Sánchez, Miguel Guzman-Merino, Christian Duran, Concepción Delgado-Sanz, Diana Gomez-Barroso, Jesus Carretero, Maria-Cristina Marinescu

## Abstract

**Background:** This work analyses the impact of different vaccination strategies on the propagation of COVID-19 within the Madrid metropolitan area starting the 27th of December 2020 and ending in the Summer of 2021. The predictions are based on simulation using EpiGraph, an agent-based COVID-19 simulator.

**Methods:** We briefly summarize the different interconnected models of EpiGraph and then we provide a comprehensive description of the vaccination model. We evaluate different vaccination strategies, and we validate the simulator by comparing the simulation results with real data from the metropolitan area of Madrid during the third wave.

**Results:** We consider the different COVID-19 propagation scenarios on a social environment consisting of the ten largest cities in the Madrid metropolitan area, with 5 million individuals. The results show that the strategy that fares best is to vaccinate the elderly first with the two doses spaced 56 days apart; this approach reduces the final infection rate and the number of deaths by an additional 6% and 3% with respect to vaccinating the elderly first at the interval between doses recommended by the vaccine producer.

**Conclusion:** Results show that prioritizing the vaccination of young individuals would significantly increase the number of deaths. On the other hand, spacing out the first and second dose by 56 days would result in a slight reduction in the number of infections and deaths. The reason is the increase in the number of vaccinated individuals at any time during the simulation.

## 1. Introduction

Immunization saves between 4 and 5 million lives annually. Its benefits extend beyond the vaccinated to include those who cannot themselves be vaccinated - small children, people with weak immune systems or those with contraindications. Resources that are saved due to fewer illnesses and hospitalizations can be invested into researching other diseases and caring for the otherwise ill [9]. At the end of 2020 the first COVID-19 vaccine batches were available in Spain and the authorities were faced with the problem of how to best schedule the different population segments for immunization considering a limited number of existing doses at this time [1]. The mitigation objectives were multiple: the immunization policies seek to reduce the number of deaths but also the number of infections (to curb transmission), of hospitalizations (to reduce the pressure on ICUs), or the incidence over specific collectives. The complexity of this problem increases with the large number of factors that have to be considered regarding the different types of vaccines - each one with different efficacies, batch sizes, and availability - as well as the surge of new virus mutations with different levels of resistance to vaccines. In this context, starting October 2020 we have been designing plausible vaccination scenarios as part of the Spanish Health Ministry task force, and we used EpiGraph [4] to simulate them and compare the outcomes in terms of COVID-19 infections, hospitalizations, and deaths. In this work we make the simulator source code and results available.

## 2 Methods

### 2.1 Simulator overview

EpiGraph consists of several different models that together reproduce the most important aspects of the simulation environment. Figure 1 shows an overview of the dataflow and simulator structure; a detailed description can be found in [2]. The *social model* reproduces the characteristics of the population that is being simulated, including the demographic data, daily activities, and interaction patterns. The experiments we report on in this paper are for the Madrid province which has about 5 million individuals, based on demographic data from the Spanish census [3]. Daily activities are structured around work/study time, leisure time, and family time. In order to realistically reproduce the social mixing, EpiGraph considers four different collectives (workers, students, the unemployed, and elderly people) as well as ten different professions. Some professions have specific contact patterns.

**Figure 1:**
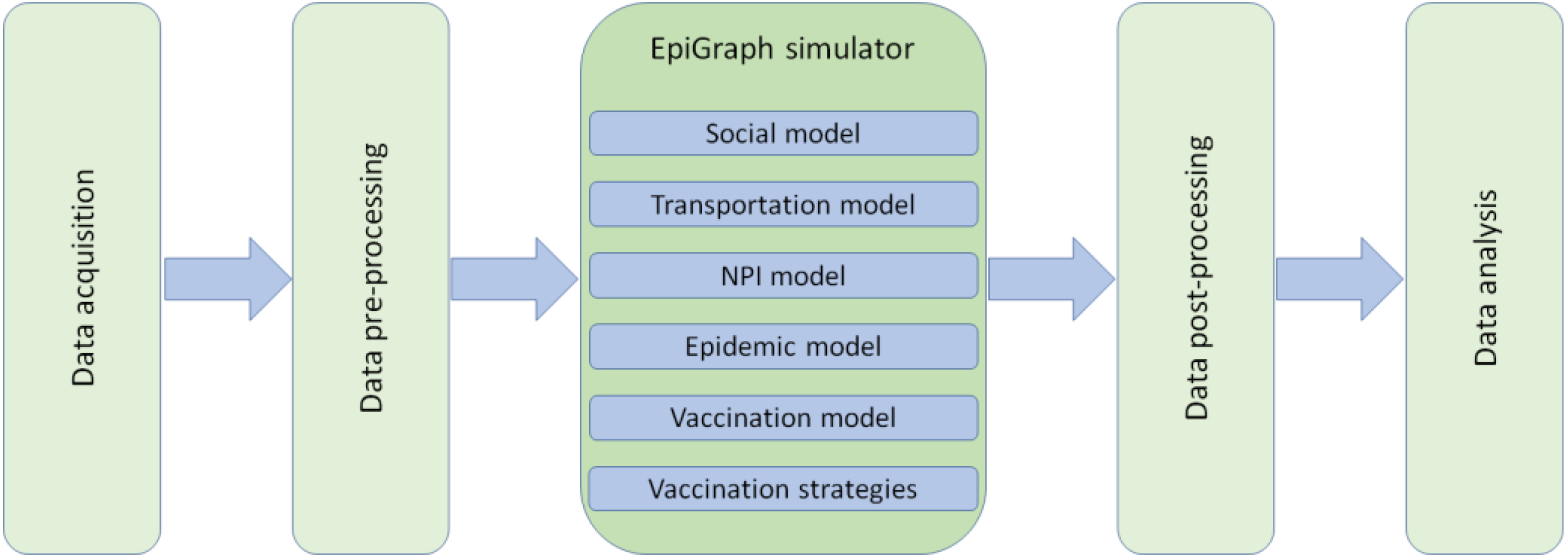
EpiGraph data flow: data acquisition and preprocessing to read and configure the input data, followed by agent-based simulation and output data post-processing and analysis

The *transportation model* considers that some individuals commute between cities. The *NPI model* reproduces the non-pharmaceutical interventions followed during the simulation period and include the use of facemasks [3], social distancing restrictions imposed by the authorities and testing of the population [4]. The *epidemic model* determines how the infection evolves in a host and specifies the probability of transmitting the virus from the infected individual to her contacts, depending on the host&s current infection stage. This probability depends on the characteristics of the individual potentially being infected, such as their age, profession or whether the individual is using a mask or is vaccinated. The epidemic model is based on a compartmental stochastic SEIR model, shown in Figure 2(left), extended to include latent, asymptomatic, dead, hospitalized, and vaccinated states. This model is independently applied to each individual. A more detailed description parameters used in the models can be found in the Supplementary Material and in [1, 4].

**Figure 2:**
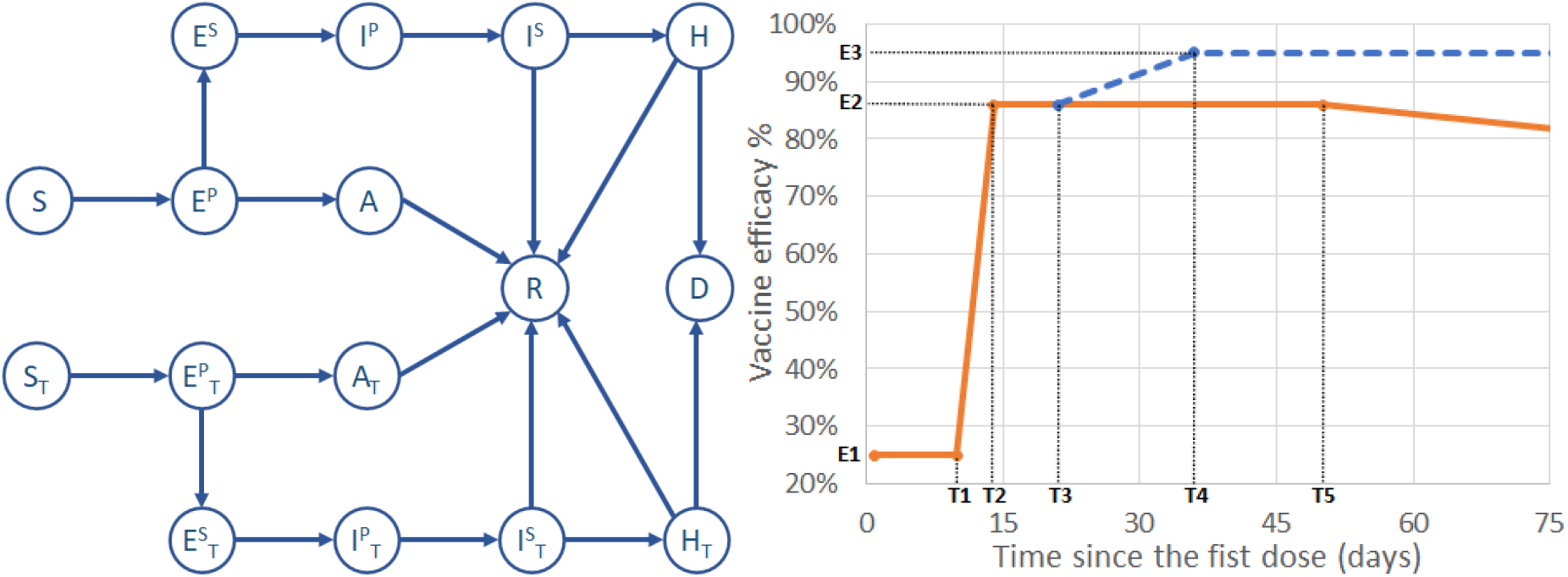
Left: Compartmental model used by EpiGraph that consists of the following states: susceptible (S), primary exposed (E^P^), secondary exposed (E^S^), asymptomatic (A), primary infected (I^P^), secondary infected (I^S^) hospitalized (H), recovered (R) and dead (D) individual. States with subindex T (ST, E^P^T, etc.) are related to vaccinated (treated) subjects. Right: Vaccination model with values corresponding to Pfizer-BioNTech vaccine. The efficacy of a single dose is displayed in a solid orange line. The second dose efficacy is represented as a dashed blue line. E1, E2 and E3 represent the minimum, first and second dose efficacies, respectively. T1 is the time when the first dose starts increasing the efficacy, T2 is the time for achieving the maximum efficacy of the first dose. T3 is the time when the second dose was applied, T4 is the time for achieving the maximum efficacy of the second dose and T5 is the time when the first dose starts decreasing its efficacy.

### 2.2 Vaccination model

This model determines the effectiveness of a vaccine for an individual according to the COVID-19 variant, vaccine type, whether it is the first or second dose (if applicable), and personal characteristics such as age or whether he/she has been previously infected with COVID-19. In the compartmental model shown in Figure 2(left) we distinguish whether the individual is vaccinated or not. A vaccinated individual is categorized as *treated*. Supplementary Material includes a description of these states as well as the transition probabilities and R0 values for each state.

We model four different vaccines in this work: Comirnaty (Pfizer-BioNTech), Spikevax (Moderna), Astra-Zeneca and Janssen (Johnson & Johnson). Figure 2(right) shows the distribution used to model the vaccine efficacy over the time. This efficacy is defined as the probability of transitioning to the Asymptomatic Treated state (AT). For example, a vaccine efficacy of 95% means that 95% of the vaccinated individuals will transition to the AT state and remain asymptomatic if they are infected, without risk of severe symptoms. The remaining 5% will transition to the E^S^T state if infected and a fraction of them will develop severe symptoms. Note that each compartmental state related to a vaccinated individual has an R0 value that is different from the case when no vaccination was applied and is dependent on the vaccine type and COVID-19 variant.

In Figure 2(right) we assume that the first dose is administered at time zero. Then, at time T1 this dose starts providing protection to the individual, which reaches the maximum efficacy value at T2. As an example, the maximum efficacy for a first dose (E2) of Pfizer-BioNTech is 52% [5]. In the figure we can observe that the first vaccine dose becomes effective after day 12. During the first 12 days [5] the vaccine does not provide any protection, while during the next 8 days the efficacy of the vaccine increases linearly to 52%. If the second dose is not applied, then the efficacy will start decreasing after time T5, reaching the minimum value E1 (25% in the figure) a year after the first dose was administered.

Note that in our model we consider that a person is naturally asymptomatic in 25% of the cases which corresponds to the probability of transitioning to the Asymptomatic state (A). The model also considers the effect of a second dose, applied at time T3^1^. In this example, the effect of the second dose starts at 52% and linearly increases to the maximum efficacy, around day 38. In this example, the maximum efficacy of E3 is 95% for Pfizer-BioNTech [5]. In this case, this maximum value is maintained for one year.

### 2.3 Vaccination strategies

The vaccination strategies reflect policies that can be adopted by health authorities worldwide and have as an objective to determine what is the most effective approach to vaccinate the population. As a general rule applied by health authorities in Spain, a candidate is subject to receiving the vaccine if he/she is in the susceptible, exposed primary, or asymptomatic states. In addition, an individual that has been infected with COVID-19 before being vaccinated will receive a single vaccine dose.

In this work we have considered five different vaccination strategies, which we describe below:

- **No vaccination**. No vaccines are administrated during the simulated time interval.
- **Elderly First (baseline)**. This strategy represents the baseline scenario which reproduces the vaccination strategy followed in Spain. For Pfizer-BioNTech and Moderna, the first group scheduled for vaccination were the elderly people living in nursing homes, their caregivers, and the front-line health professionals. The remaining health workers were vaccinated next, followed by the general population in age-decreasing order. For Astra-Zeneca the target was people between 18 and 56 years old before March 23th; 18 and 65 years old between March 23th and April 9th; and 60 and 69 years old after April 9th. Priority was given to elderly caregivers and health, security forces, and education professionals. In the simulations the Janssen vaccine was only delivered to people between 50 and 70 years old.
- **Young First**. This policy prioritizes individuals following an age-increasing order for the Pfizer-BioNTech and Moderna vaccines. With this policy, the first social group that receives the vaccine are teenagers, followed by people between 20 and 29 years old, and so on. The idea for this strategy is to limit the impact of transmission via groups that have the most social contacts within the entire population and tend to have the least noticeable symptoms. No specific professions are prioritized. This strategy stays unchanged compared to the “Elderly First” for Astra-Zeneca and Janssen, a decision taken by the Spanish authorities.
- **Elderly First, 56 days between doses (56D)**. This strategy represents a variation of the Elderly First strategy in which the first and second doses are separated by 56 days (instead of 21) for the Pfizer-BioNTech and Moderna vaccines. For Astra-Zeneca and Janssen the strategy is the same as the “Elderly First”. This is a scenario that the authorities tested to assess whether it could protect a larger fraction of the population from extreme symptoms that could lead to hospitalization and death.
- **Elderly First, 2 doses already infected (2DI)**. This strategy represents a variation of the Elderly First strategy in which individuals who have been previously infected with COVID-19 also receive two doses of Pfizer-BioNTech or Moderna vaccines. The strategy for the rest of the population is the same as in the Elderly First.

## 3 Results

### 3.1 Model validation

EpiGraph was validated using the Madrid province (Spain) as a simulation scenario; this area mostly consists of the metropolitan area of Madrid which includes the city of Madrid and the following cities: Alcalá de Henares, Alcobendas, Alcorcón, Fuenlabrada, Getafe, Leganés, Móstoles and Parla, for a total of 5,018,241 inhabitants. EpiGraph was executed on Tirant supercomputer at the University of Valencia. The simulations for all scenarios start on December 27th, 2020, which corresponds to the initiation of the COVID-19 vaccination campaign in Spain. The simulation time span is 190 days. Fig. 3(left) shows the aggregated number of infected individuals, with real data in red [20] and simulated data in blue. Each city includes demographic information which corresponds to the Madrid province and reflects the real population pyramid, job sector distribution, number of family members per household, etc. EpiGraph uses stochastic processes to perform the simulations, which may result in differences between the results in every run. In order to quantify the deviation in the results, we have repeated the same simulation 10 times obtaining a median number of notified daily cases of 3,111 for the simulation timeline (from December 27th 2020 and July 5th 2021). This value is similar to the average number of reported of 3,255 cases during the same period.

**Figure 3:**
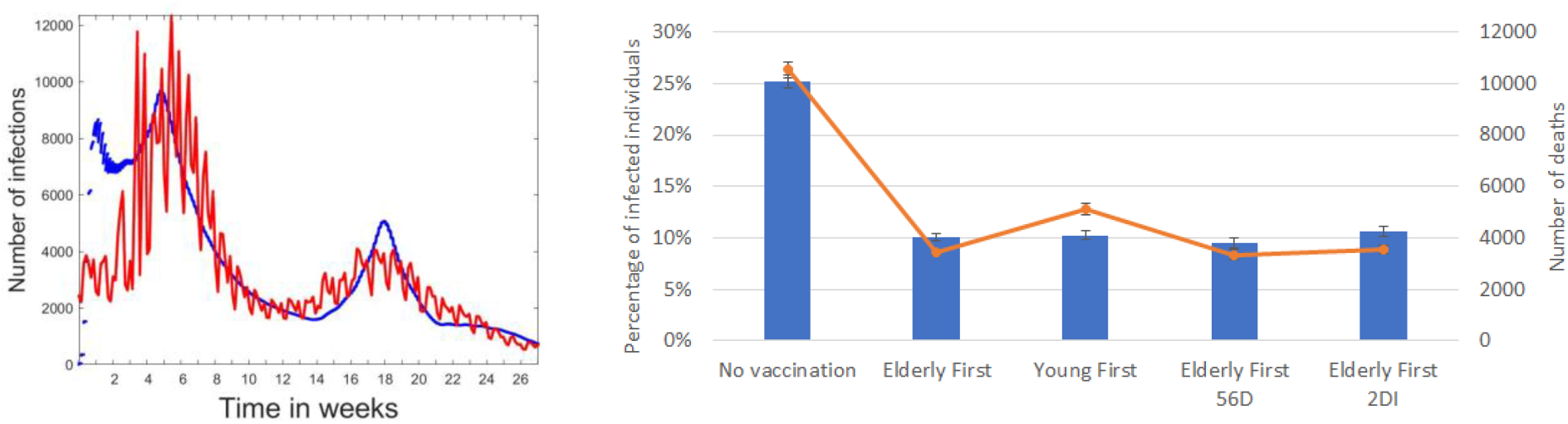
On the left, results for model validation. These results include the aggregated number of infected individuals for the area under study. Real and simulated data are shown in red and blue, respectively. The simulated curve corresponds to the baseline scenario. On the right, simulation results that include the percentage of infected population in blue bars and the number of deaths in orange, for each one of the vaccination strategies.

### 3.2 Vaccination scenarios

Figure 3(right) shows, for each vaccination strategy, the percentage of infected individuals from the overall population and the number of deaths at the end of the simulation. Each result corresponds to the median value of ten different simulation runs and includes an error bar that represents the standard deviation. In addition to face masks, all scenarios implement several social distancing restrictions that reduce the capacity in restaurants and social gatherings. Figures 4 and 5 show the number of infections and deaths broken down by groups. From Figure 3(right) we can observe that the no-vaccination scenario results in nearly 150% more infections and 200% more deaths than any of the vaccination scenarios. According to our model, Young Firsts results in a similar number of total infections and about 48% more deaths than the baseline approach (Elderly First). The reason is that one of the most vulnerable groups - the elderly, are not vaccinated until the end of the simulation. In Figure 5 we can observe that most of the deaths are among elderly people. Spacing out the first and second doses by 56 days increases the number of people vaccinated at any time, although they may only have received one dose. According to our model, this approach reduces the final infection rate and the number of deaths by 6% and 3% with respect to the baseline. Lastly, the Elderly First 2DI strategy decreases vaccination coverage, which results in a net increase of 5% and 1% in the number of infections and deaths, respectively - compared to the baseline.

**Figure 4:**
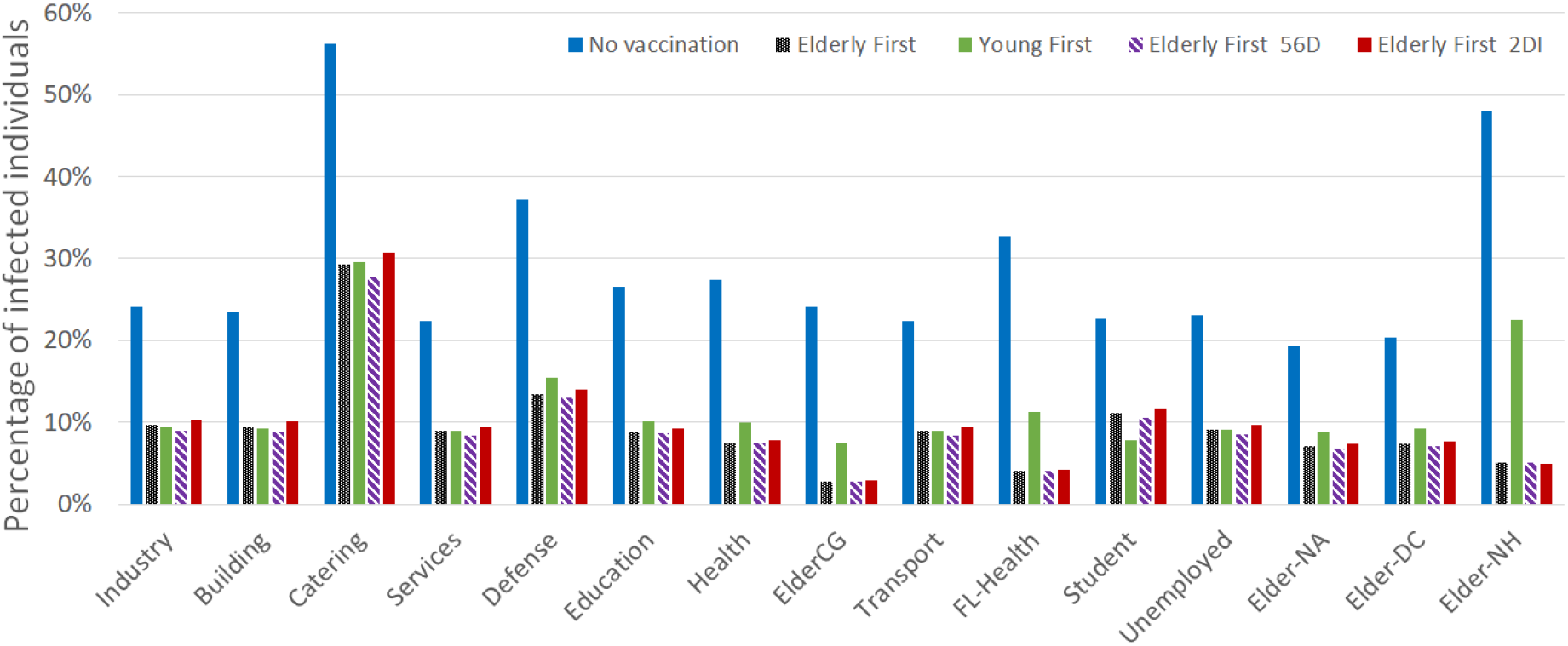
Percentage of infected population at the end of the simulation, by groups. The acronyms stand for: ElderCG - caregiver for elderly people; Health - non-front-line health professionals; FL-Health - front-line health professionals; Elder-NA - elderly that live by themselves; Elder-DC - elderly attended in daily centers; and Elder-NH - elderly that live in nursing homes.

**Figure 5:**
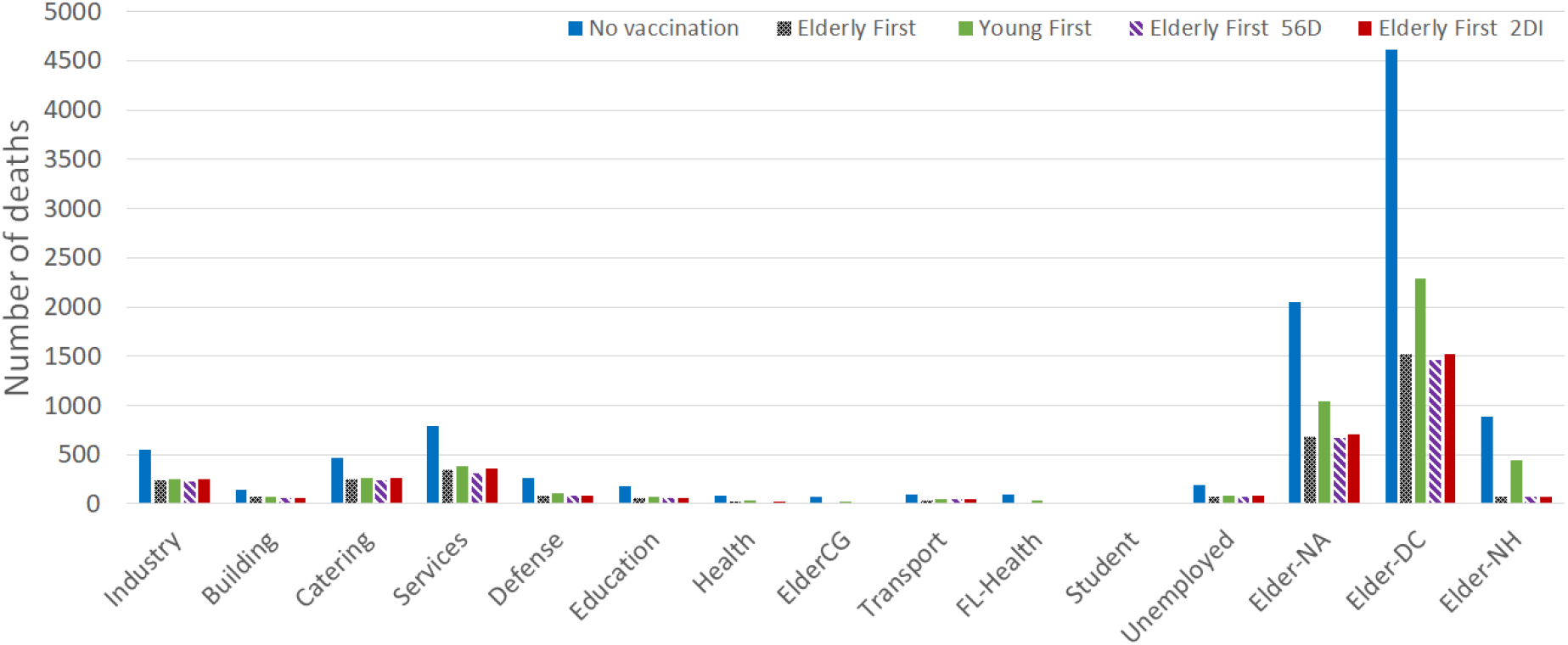
Number of deaths at the end of the simulation, broken down by groups. The acronyms are identical to those in Figure 4.

## 4 Discussion

The nature of EpiGraph as an agent-based model gives it the power to directly represent characteristics of the population under study, as well as the possibility to model the effect of other factors, e.g. NPIs or vaccination strategies, on the individuals in a population. This allows us to (relatively) easily add individual attributes that are relevant, model different interventions, customize and refine them, and observe their effects for the different segments of population. According to the results obtained in this work, spacing out the first and second dose by 56 days would result in a slight reduction in the number of infections and deaths. The reason is the increase in the number of vaccinated individuals at any time during the simulation, although some may only have the first dose. Prioritizing the vaccination of young individuals would significantly increase the number of deaths.

In [6], the authors use a deterministic SEIR framework to model the propagation of the virus and the effect of non-pharmaceutical interventions (social distancing mandates and mask use) until Spring of 2021. Some of the limitations of this approach are the absence of age structure and the assumption of a well-mixed population. Covasim [7] includes demographic information about age structure and population size. Different from our work, the contacts are not based on existing patterns; scalability issues are partly sidestepped by dynamic scaling. Vaccines are modelled by adjusting individuals’ susceptibility to infection and probability of developing symptoms after being infected; both of these modifications affect the overall probability of progressing to severe disease and death. However, some features we consider in EpiGraph (like vaccine effectiveness across variants) are not currently implemented in Covasim. Modelling social mixing a crucial factor for obtaining realistic simulations. In [8, 10, 11] different ways for refining the social interactions are considered. In EpiGraph the social mixing modelling is carried out using Facebook and Enron contact networks and individual contact matrices.

[12] compares five age-stratified prioritization strategies in terms of cumulative incidence, mortality, and years of life lost. Some limitations have to do with using pre-pandemic contact matrices, not incorporating nonpharmaceutical interventions, and only considering variation in disease severity and risk by age - although contact rates, and thus infection potential, vary greatly not only by occupation and age. Results show, like in our work, that people aged 60 years and older should be prioritized to minimize deaths. In [13] the authors use a mathematical model paired with optimization algorithms to determine the optimal use of vaccine for different combinations of vaccine effectiveness and number of doses available under a wide variety of scenarios; the optimal allocation strategies were computed using age as the sole risk factor. This work obtains similar conclusions than in our work, that is, for low vaccine effectiveness, the best option for reducing deaths is to allocate vaccines to older age-groups first.

The work [14] uses a mathematical model to assess the optimal allocation of a limited vaccine supply in the United States across groups differentiated by age and essential worker status, which constrains opportunities for social distancing. The authors show how optimal prioritization is sensitive to several factors including vaccine effectiveness and supply, and rate of transmission. [15] uses the agent-based infectious disease modelling tool CovidSIMVL to explore outcomes of 2-dose vaccination regimens and a third “Hybrid” policy that reflects ranges of expected levels of protection according to Pfizer and Moderna, but with a 35-day separation between first and second dose instead of the shorter recommended period. Unlike in our work, agents here were considered homogeneous and vaccination strategies were not stratified (e.g., by age ranges) nor sequenced in order to best manage risk on the basis of considerations of population-level transmission risk and on the basis of considerations of equity.

[16] proposes a multi-scale agent-based model to investigate the infectious disease propagation between cities and within a city using the knowledge from person-to-person transmission. This is a way to reduce the degree of freedom of the model as follows: at micro scale, an agent represents a person while at the meso scale an agent refers to hundreds of individuals. Actual data on traffic patterns and demographic parameters are adopted however, unlike in our work, no age stratification is considered in the vaccination. In [17] a study of how Genetic Algorithms (GA) can be applied to an ABM in order to provide parameter estimates for administering the vaccine to groups of people. One of the important variables defines the working condition of agents, including categories of interest in the contrast of Covid-19 pandemic, i.e. hospital healthcare operators, nursing home healthcare operators, teachers, students, workers, and fragile workers. [18] proposes a second dose delay strategy for people below 65 years old. Like in our work, this strategy shows benefits reducing the number of infections.

The main limitations of EpIGraph are that we do not consider attributes such as previous pathologies that we now know that may come into play when we evaluate the risk of developing COVID-19 severe symptoms. In the transportation model the movement of individuals between cities depends only on the distance between the cities and the population size. In our experiments we only model the largest urban regions in the Madrid metropolitan area; we could incorporate smaller cities and towns to the simulation, including rural regions.

In conclusion, results show that prioritizing the vaccination of young individuals would significantly increase the number of deaths. On the other hand, spacing out the first and second dose by 56 days would result in a slight reduction in the number of infections and deaths. Similar results have been obtained in other published works. This study goes further to obtain refined results by age and profession and adds a detailed and realistic vaccination model. Our immediate plans include modelling and evaluating the effect of a third vaccination dose and simulating vaccination scenarios for the entire area of Spain and the many variants that have appeared since the beginning of the vaccination campaign (including potential vaccine resistant variants). Finally, these results helped health authorities to adjust the COVID vaccination strategy in Spain to reach better results. These tools, which allow us to adapt to changes and predict future situations, are essential to achieve the best health decisions with the most efficient use of resources.

## Data Availability

All data produced in the present study are available upon reasonable request to the authors

## Ethics approval and consent to participate

Not applicable. This work uses public data and performs simulations not involving experiments with humans nor animals.

## Authorship and contributorship

David E. Singh, Miguel Guzman-Merino and Christian Duran designed and implemented EpiGraph simulator. David E. Singh, Carmen Olmedo Lucerón and Aurora Limia Sánchez conceived and designed the experiments. David E. Singh, Miguel Guzman-Merino and Christian Duran processed the input data and ran the experiments. David E. Singh, Carmen Olmedo Lucerón, Aurora Limia Sánchez and Maria-Cristina Marinescu wrote the paper. David E. Singh, Concepción Delgado-Sanz, Diana Gomez-Barroso, Carmen Olmedo Lucerón, Aurora Limia Sánchez, Maria-Cristina Marinescu and Jesus Carretero provided insights on the validity of our assumptions, recommended additional related work, and contrasted the results with their own findings. All authors review the different manuscript drafts and approved the final version for submission.

## Footnotes

1 Note that T3 may be bigger than T5. In this case, the vaccine efficacy is also increased by the second dose until reaching the maximum value of E3.

