## Supplemental data for "Evaluation of Vaccination Strategies for the metropolitan area of Madrid"

[1] Universidad Carlos III de Madrid, Leganes, Spain.

[2] Vaccine area. Spanish Health Ministry. Spain

[3] CIBER en Epidemiología y Salud Pública (CIBERESP), Madrid, Spain; National Centre for Epidemiology, Carlos III Institute of Health, Madrid, Spain.

[4] Barcelona Supercomputing Center, Barcelona, Spain.

*This work has been supported by the Carlos III Institute of Health under the project grant 2020/00183/001, the project grant BCV-2021-1-0011, of the Spanish Supercomputing Network (RES) and the European Union's Horizon 2020 JTI-EuroHPC research and innovation program under grant agreement No 956748. The role of all study sponsors was limited to financial support and did not imply participation of any kind in the study and collection, analysis, and interpretation of data, nor in the writing of the manuscript.

### 1. Social model

The following tables show the different parameters used to configure the social model used by EpiGraph in our experiments. It is important to highlight that these parameters are related to the demographic and social conditions of each of the considered regions of Spain. In order to synthesize our results, we show the input parameters used for the province of Madrid. The data was collected from the Spanish National Statistics Institute (INE) [1]. The population pyramid (not shown in tables) was also collected from the INE for each Spanish province.

Table 1 shows the percentage distribution of each collective and the sizes of the groups considered for each collective. In Table 2 the work collectives are broken down by profession and include the industry, construction, catering services, security, education, health, elderly care, and transportation. Note that some of the professions have specific contact patterns, which are considered in the social model. More specifically, education and elderly care include static contacts with students and elderly people at nursing home, respectively. For catering, security, and health we consider dynamic contacts. Health professionals are also divided into front-line and non-front-line workers. Each one of these two sub-collectives have different types of dynamic contacts. In Table 3 the elderly collective is broken down by sub-collectives: elderly people at home, in day-care centers, and in nursing homes. Table 4 illustrates the family size distribution used in our simulation. Note that this distribution is also different for each province. Table 5 shows the list of parameters used to model the individual (i.e. agent). We distinguish between static parameters - with constant values- and dynamic parameters - which may change during the simulation. The table also indicates whether the parameter is used during the simulation.

| | | | | | |
|---|---|---|---|---|---|
| School groups | | | | | |
| MinAge | 0 | MaxAge | 19 | Percentage | 0.1757% |
| MinSize | 40 | MaxSize | 200 | Percentage males | 0.5108% |
| Work groups | | | | | |
| MinAge | 20 | MaxAge | 64 | Percentage | 0.5179% |
| MinSize | 20 | MaxSize | 1000 | Percentage males | 0.4770% |
| Stay-at-home, informal meetups groups | | | | | |
| MinAge | 20 | MaxAge | 64 | Percentage | 0.1194% |
| MinSize | 1 | MaxSize | 10 | Percentage males | 0.4770% |
| Elder, informal meetups groups | | | | | |
| MinAge | 65 | MaxAge | 100 | Percentage | 0.1870% |
| MinSize | 25 | MaxSize | 50 | Percentage males | 0.3905% |

*Table 1: Social group distribution for the cities of Madrid province.*

| | Industry | Construction | Catering | Services | Security | Edu. | Health | Elderly-CG | Transport |
|---|---|---|---|---|---|---|---|---|---|
| | 30.80% | 6.50% | 8.80% | 24.00% | 7.40% | 7.50% | 6.40% | 3.30% | 5.30% |
| $Size_{min}$ | 1 | 1 | 1 | 1 | 10 | 6 | 10 | 5 | 1 |
| $Size_{max}$ | 30 | 20 | 12 | 8 | 50 | 30 | 30 | 25 | 8 |

*Table 2: Work collective breakdown in professions. Edu. and Elderly-CG stands for education professionals and elderly caregivers, respectively. The percentages are the fraction of each profession among the worker collective.* $Size_{min}$ *and* $Size_{max}$ *denote the minimum and maximum sizes of each specific collective. A normal distribution between these two values has been used for setting each group size.*

| | Elderly at home | Elderly at day-care centre | Elderly at nursing home |
|---|---|---|---|
| | 50.6% | 46.3% | 3.1% |
| $Size_{min}$ | 4 | 10 | 10 |
| $Size_{max}$ | 10 | 30 | 40 |

*Table 3: Elderly collective breakdown in classes. Elderly at home represents the elderly people that live at home and participate in day centers (in our simulations, according to the existing conditions in Spain, day centers were closed during the simulation period so this collective was merged with the elderly-at-home collective). The percentages are the fraction of each class among this collective.* $Size_{min}$ *and* $Size_{max}$ *denote the minimum and maximum sizes of each specific collective. A normal distribution between these two values has been used for setting each group size.*

| Number of members in a family | | | | | |
|---|---|---|---|---|---|
| 1 member: | 25.50 % | 2 members: | 30.40 % | 3 members: | 20.90% |
| 4 members: | 17.70 % | 5 members: | 5.50 % | | |

*Table 4: Family size distribution for the cities of the Madrid metropolitan area.*

| *Parameter* | Type | Description | Used |
|---|---|---|---|
| Age | Static | Individual age | Yes |
| Gender | Static | Male or female | No |
| Etnic group | Static | White, black, latino, asian, american indian, others | No |
| Occupation | Static | Student, worker, elderly people or unemployed | Yes |
| Occupation group | Static | Profession. See Table 2 | Yes |
| Work on Saturday | Static | If true, the individual works on Saturdays | Yes |
| Health condition | Dynamic | Factors than can increase the risk of severity synonym | No |
| Mask use | Dynamic | Mask use, type of mask | Yes |
| Quarantined | Dynamic | Isolation | Yes |
| Vaccination type | Dynamic | Vaccine type: Pfizer-Biontech, Moderna, Astra-Zeneca or Janssen. | Yes |
| Vaccination t1 | Dynamic | Vaccination time of the first dose | Yes |
| Vaccination t2 | Dynamic | Vaccination time of the second dose | Yes |
| Infection stage | Dynamic | If infectious, the infection stage related to the individual. See Table 7 | Yes |
| COVID-19 variant | Dynamic | COVID-19 variant: Wuhan, British, E484K or Delta | Yes |
| Infection t1 | Dynamic | Infection start time | Yes |
| Infection t2 | Dynamic | Infection end time | Yes |
| Sick time | Dynamic | Time that the individual was on bed because of the illness | Yes |
| Seroprevalence | Dynamic | Prevalence to SARS-COV-2 | Yes |
| Sequels | Dynamic | Infection sequels | No |
| Test type | Dynamic | Testing method used | Yes |
| Test time | Dynamic | Testing time | Yes |
| Extra daily tests | Static | Extra PCR tests in the strategies | Yes |
| Quarantine breakers | Static | Percentage of individuals that break quarantine time | Yes |
| Test window | Static | Days for testing the same individual consecutively | Yes |

*Table 5: List of parameters used to model the agent. The column labelled $Type$ indicates whether the parameter is static or dynamic, i.e. it has a constant value during the simulation or its value is may change. All these parameters are implemented but only the used ones determine the infection outcome.*

### 2. Epidemic model

In Figure 2(left) of the main article, when infected, the individual transitions from Susceptible Treated ($S_T$) to Exposed Primary Treated ($E^P_T$) and then to one of two possible states, as follows. In case of vaccine failure, the individual transitions to the Exposed Secondary Treated ($E^S_T$) state and then to Infected Treated Primary and Secondary states ($I^P_T$ and $I^S_T$, respectively). In these states the individual is at risk of being Hospitalized ($H_T$) or dying(D). If, on the other hand, there is no vaccination failure, then the individual transitions to the Asymptomatic Treated state ($A_T$). Note that in this state the individual may spread the disease to a lesser extent that for a non-vaccinated infected individual but will not experience any health condition herself. Tables 6 and 7 show the $R_0$ values and transition probabilities for each compartment state considered in the Epidemic model.

| Compartment state | $R_0$ values | | Probability | |
|---|---|---|---|---|
| $E^P$ | $R_0^{EP}$ | 0 | $P^A$ | 25% |
| $E^S$ | $R_0^{ES}$ | 1.42 | | 100% |
| $A$ | $R_0^A$ | 1.42 | | 100% |
| $I^P$ | $R_0^{IP}$ | 4.5 | $P^{IS}$ | 100% |
| $I^S$ | $R_0^{IS}$ | 3.38 | $P^H$ | Table 7 |
| $I^{SV}$ | $R_0^{ISV}$ | N/A | | 100% |
| $H$ | $R_0^H$ | 0.34 | $P^D$ | Table 7 |
| $E^P{}_T$ | $R_0^{EP}$ | 0 | $P^{AT}$ | 25% |
| $E^S{}_T$ | $R_0^{ES}$ | 0 or 1.42 | | 100% |
| $A_T$ | $R_0^A$ | 1.1 or 1.42 | | 100% |
| $I^P T$ | $R_0^{IP}$ | 0 or 4.5 | $P^{IS}$ | 100% |
| $I^S{}_T$ | $R_0^{IS}$ | 0 or 3.38 | $P^H$ | Table 7 |
| $H_T$ | $R_0^H$ | 0 or 0.34 | $P^D$ | Table 7 |

*Table 6: $R_0$ Values and transition probabilities for each compartment state. In this work we have not considered the use of antivirals, thus $I_V{}^S$ state is not reached and the associated $R^{ISV}{}_0$ value is not applicable. $E^S$ and $A$ states do not have a related transition probability because there is only a destination state. $P^{AT}$ represents the transition to asymptomatic for vaccinated individuals. This probability is vaccination-dependent.*

| | Age interval | | | | | | | | |
|---|---|---|---|---|---|---|---|---|---|
| | < 10 | 10-19 | 20-29 | 30-39 | 40-49 | 50-59 | 60-69 | 70-79 | ≥80 |
| $P^H$ | 0.4% | 0.4% | 3.4% | 9.0% | 19.6% | 31.4% | 40.8% | 49.8% | 45.2% |
| $P^D$ | 0.0% | 0.4% | 0.8% | 0.8% | 1.2% | 2.0% | 4.7% | 12.2% | 30.0% |

*Table 7: Values of $P^H$ and $P^D$ are based on age. $P^H$ is the probability an infected person has of becoming hospitalized and $P^D$ is the probability a hospitalized person (a fraction of the total infected) has of dying.*

### 3. Vaccination model

Table 8 shows the parameters used in the vaccination model. E1, E2 and E3 represent the minimum, first and second dose efficacies, respectively. T1 is the time when the first dose starts increasing the efficacy, T2 is the time for achieving the maximum efficacy of the first dose. T3 is the time when the second dose was applied, T4 is the time for achieving the maximum efficacy of the second dose and T5 is the time when the first dose starts decreasing its efficacy. Table 9 shows the daily doses per 100,000 habitants for each vaccine type.

| Vaccine | *E1* | *E2* | *E3* | *T1* | *T2* | *T3* | *T4* | *T5* |
|---|---|---|---|---|---|---|---|---|
| Comirnaty | 0.25 | 0.86 | 0.95 | 7 | 14 | 21 | 28 | 365 |
| Spikevax | 0.25 | 0.86 | 0.95 | 7 | 14 | 28 | 35 | 365 |
| Astra-Zeneca | 0.25 | 0.6 | 0.7 | 7 | 14 | 84 | 98 | 365 |
| Janssen | 0.25 | 0.8 | n/a | 7 | 28 | n/a | n/a | 365 |

*Table 8: Parameters used in the vaccine model.*

| Month | **Moderna** | **Comirnaty** | **AstraZeneca** | **Janssen** |
|---|---|---|---|---|
| January | 24 | 152 | 0 | 0 |
| February | 24 | 152 | 132 | 0 |
| March | 24 | 152 | 102 | 0 |
| April | 133 | 341 | 105 | 133 |
| May | 133 | 344 | 105 | 133 |
| June | 133 | 781 | 105 | 133 |
| July | 144 | 502 | 232 | 289 |
| August | 144 | 502 | 232 | 289 |
| September | 144 | 502 | 232 | 289 |

*Table 9: Considered daily doses per 100,000 habitants for each vaccine type.*

### 4. Simulation configuration

The simulation starts with an initially infected population (per city) of 0.6%, a number that corresponds to the officially reported cases at the end of December 2020. EpiGraph is calibrated only once for the baseline scenario over the entire simulation period; this is the scenario that reproduces the actual vaccination strategy that has been applied during the simulation period. The initial conditions include prevalence values at simulation start time, i.e. the percentage of the population that had already recovered from COVID-19 before the start of the third wave in Spain. These values are 11% for workers, 9.1% for students, 8.6% for unemployed and 1.01% for elderly people; we assume that these individuals have become immune to COVID-19 and they will not become re-infected during the entire simulation time. The parameters related to the epidemiological and NPI models are taken to be the same for all the cities under study and were not involved in the calibration process. In terms of NPIs, we reproduce the social distancing measures that were applied in the Madrid metropolitan area during the simulation period. As a result, all individuals use face masks at work, school, and during leisure time, but not when they are at home.
